# Integrated Framework for the Optimal Determination of Diagnostic Cut-off Points through Empirical Interpolation, Logistic Modeling Optimized by Dual Annealing, and Combinatorial Optimization with ThresholdXpert: Application to Hepatocellular Carcinoma

**DOI:** 10.64898/2026.02.19.26346674

**Authors:** Roberto Reinosa Fernández

## Abstract

**Introduction:** The precise determination of diagnostic cut-off points is essential for the development of multimarker panels in oncology. In previous work on pulmonary nodules, it was observed that the standard two-parameter logistic fit could be insufficient for biomarkers with asymmetric distributions. Furthermore, the calculation of empirical cut-off points based on graphical visualization presented limitations in precision and reproducibility.

**Objective:** This study presents a methodological advancement in the data analysis phase (Stage 1), introducing new Python algorithms for the direct analytical calculation of empirical intersections and robust mathematical modeling using Dual Annealing with both two-parameter and four-parameter logistic functions. This improved methodology feeds into the ThresholdXpert 1.0 software tool for combinatorial optimization of biomarker panels (Stage 2), and is applied here to the diagnostic challenge of hepatocellular carcinoma (HCC).

**Methods:** The methodology was first validated by re-analyzing a dataset of patients with pulmonary nodules (N=895). It was subsequently applied to an HCC dataset derived from the cohort of Jang et al. (208 HCC, 193 cirrhosis, 401 total), randomly divided into a training set (280) and an independent test set (121). Scripts were developed to compare the previous two-parameter logistic fit with the new two- and four-parameter logistic models. Finally, ThresholdXpert 1.0 was used for multimarker panel optimization.

**Results:** The integration of empirical calculation, logistic modeling, and combinatorial optimization through ThresholdXpert 1.0 provides a robust and coherent framework for the development of multimarker diagnostic panels. The four-parameter logistic model provided additional validation without substantially modifying cut-off values for most biomarkers, confirming the stability of the approach while offering greater flexibility for complex distributions. When applied to hepatocellular carcinoma, the framework identified a molecular panel composed of AFP, PIVKA-II, OPN, and DKK-1 with sensitivity of 0.77 and specificity of 0.72, and an optimized panel incorporating inverse MELD that achieved the best overall balance (sensitivity 0.73, specificity 0.75) in independent external validation. These results demonstrate the potential of this approach as a generalizable tool for the optimized design of binary diagnostic systems in oncology.

**Conclusion:** The integration of complementary mathematical modeling enhances the capability of ThresholdXpert 1.0 to identify robust diagnostic panels, as in some cases a single biomarker may outperform biomarker combinations, and vice versa. This approach enabled the integration of molecular biomarkers and clinical variables under a unified mathematical framework.

## 1. Introduction

Accurate risk stratification in patients with suspected hepatocellular carcinoma (HCC) is critical, particularly in the context of underlying liver cirrhosis [1]. Although alpha-fetoprotein (AFP) continues to be used, its individual performance is suboptimal, with sensitivities often not exceeding 60% [2]. Recent studies, such as that of Jang et al. [2], have proposed new combinations of biomarkers (PIVKA-II, Osteopontin [OPN], Dickkopf-1 [DKK-1]); however, these frequently rely on static cut-off values derived from the entire cohort, which may lead to overfitted models that fail in real-world clinical practice.

In previous research on pulmonary nodules [3], a two-stage workflow was established using the ThresholdXpert 1.0 tool. However, during its extensive application, areas for improvement were identified in Stage 1 (Individual Characterization): classical gradient descent methods may become trapped in local minima, and the simple two-parameter logistic model does not always capture the biological complexity of certain asymmetric biomarkers [4].

This study introduces improvements to the mathematical methodology prior to the use of the software:

1. **Direct Empirical Calculation:** A new algorithm based on numerical interpolation was developed to determine with precision the intersection point between sensitivity and specificity curves, eliminating the subjectivity associated with visual inspection. In previous approaches, graphical representation occasionally required the exclusion of a small number of extreme values to improve visualization, which could produce slight variations in the estimated sensitivity and specificity values [3].
2. **Advanced Stochastic Optimization:** Implementation of Dual Simulated Annealing for curve fitting. This improvement was introduced for both the two-parameter and four-parameter logistic models.
3. **Four-Parameter Logistic Modeling:** Inclusion of four-parameter logistic models to fit complex biological curves that do not asymptotically reach 0 or 1 [4].

These improvements were first validated using the original lung cancer dataset to demonstrate methodological consistency and were subsequently applied to develop new HCC panels using rigorous external validation.

## 2. Materials and Methods

### 2.1. Datasets and Preprocessing

The study used two distinct data sources:

1. **Pulmonary Nodules (Methodological Validation):** The dataset from Qiu et al. (N=895) [5], previously used in earlier work, was reused to compare thresholds obtained with the original methodology versus the new methodology.
2. **Hepatocellular Carcinoma (Clinical Application):** Data were derived from the study by Jang et al. [2], including 401 patients (208 HCC, 193 cirrhosis controls). A random split was performed, generating a training set (N=280) for modeling and optimization, and an independent test set (N=121) for blinded validation. Seven variables were analyzed: Age, CTP Score, MELD (Model for End-Stage Liver Disease) [6], AFP, PIVKA-II, OPN, and DKK-1. Both sets were balanced, with approximately 50% HCC and 50% liver cirrhosis.

**Note on MELD:** The MELD variable was analyzed both in its direct form and inverse form (1/MELD), under the hypothesis that preserved liver function could act as a positive discriminator for HCC compared to advanced cirrhosis.

### 2.2. Stage 1: Mathematical Modeling (Python Scripts)

The original scripts were replaced with new Python implementations.

- **Empirical model:** The determination of the optimal cut-off point was improved by calculating the intersection of the interpolated sensitivity and specificity curves.
- **Curve fitting:** The four-parameter logistic model was introduced:

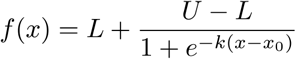

where is the lower asymptote, is the upper asymptote, is the slope, and is the inflection point.
- **Logistic model optimization:** The two-parameter logistic model was improved so that its constants were estimated using the Dual Annealing algorithm [7], as was also implemented for the four-parameter logistic model.

### 2.3. Stage 2: Optimization with ThresholdXpert 1.0

The ThresholdXpert 1.0 software was used for combinatorial search and optimization of thresholds in the training set. The primary objective of this stage was to identify combinations of variables that maximized overall diagnostic performance, seeking the best possible balance between sensitivity and specificity.

Although ThresholdXpert 1.0 is primarily designed for the development and evaluation of multimarker panels, in this study it was also used complementarily to evaluate individual markers. This provided additional validation against the mathematical models from Stage 1.

During the stochastic optimization process, fluctuations in the proposed thresholds were monitored across multiple executions. It was qualitatively observed that certain variables exhibited pronounced oscillations without providing significant improvements in overall model performance. While these fluctuations served as a useful indicator of the mathematical “noise” contributed by some variables, the final panel selection was primarily based on predictive performance (sensitivity/specificity) and clinical coherence.

## 3. Results

### 3.1. Validation of the Methodological Improvement: Re-analysis of the Pulmonary Nodule Dataset

To confirm the robustness of the new algorithms, the Pulmonary Nodule dataset was re-evaluated. Tables 1 and 2 show a comparison between the new code used to calculate the optimal cut-off point (where sensitivity equals specificity) and the previous implementation. The results were virtually identical, with the additional advantage that manual visual estimation from the graph was no longer required.

**Table 1:** Comparison of the old vs. new source code for calculating the empirical cut-off in Pulmonary Nodules (Training Set). Only a single Sen/Spe value is shown because sensitivity and specificity are equal at that point, thus having the same value.

| Variable | Old |  | New |  |
| --- | --- | --- | --- | --- |
|  | Cut-off | Training (Sen/Spe) | Cut-off | Training (Sen/Spe) |
| Age | 59.11 | 0.58 | 59.34 | 0.59 |
| BMI | 24.96 | 0.53 | 24.96 | 0.53 |
| Nodule Diameter | 14.72 | 0.72 | 14.72 | 0.72 |
| CT Value | -425.45 | 0.82 | -425.34 | 0.82 |
| CTR | 0.23 | 0.82 | 0.23 | 0.82 |
| CEA | 2.22 | 0.58 | 2.28 | 0.59 |
| CYFRA21-1 | 1.98 | 0.55 | 2 | 0.55 |
| SCC | 0.93 | 0.51 | 0.95 | 0.52 |
| ProGRP | 42.18 | 0.49 | 42.35 | 0.49 |
| NSE | 19.14 | 0.49 | 19.18 | 0.49 |

**Table 2:**
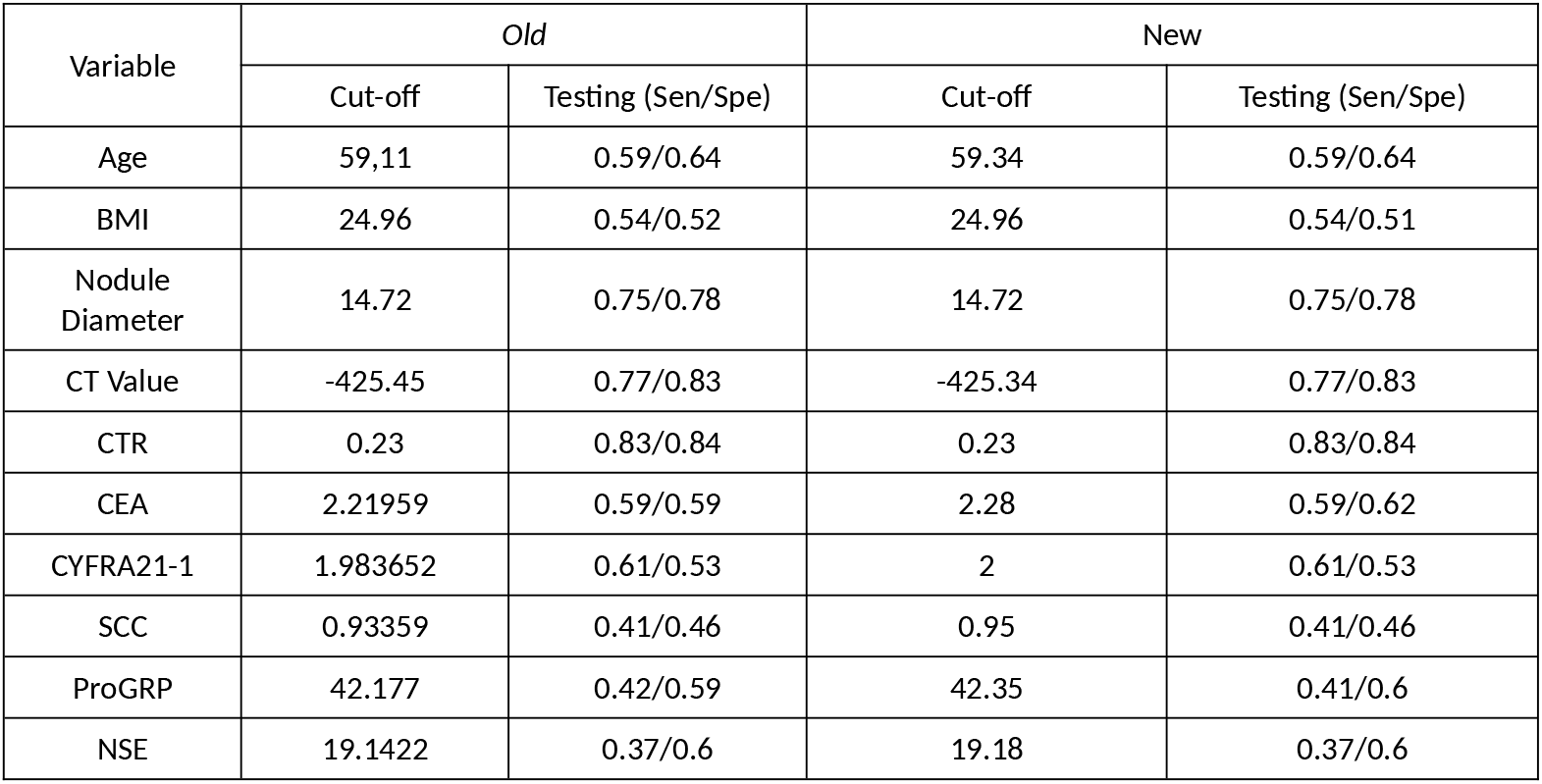
Comparison of sensitivity and specificity of the thresholds obtained with the new source code for the empirical model vs. the original ones (Test Set Validation).

In Table 3, the new methodology (two- and four-parameter logistic models combined with Dual Annealing) replicated with high precision the thresholds that previously required manual adjustment. One notable result was the calculation of the CTR threshold, where the previous methodology produced a value of 0.18, compared to 0.22 for the new two-parameter model and 0.20 for the four-parameter model. These new cut-off values are consistent with those obtained using ThresholdXpert 1.0 and with the empirical calculation.

**Table 3:** Comparison of Methodologies in Pulmonary Nodules: Old Method vs. New Models. Old 2-parameter logistic fit vs. new 2-parameter logistic fit with dual annealing and 4-parameter logistic fit with dual annealing. (Test Set)

| Variable | Old 2-parameter Cut-off | Sen/Spe | New 2-parameter Cut-off (Analytical) | Sen/Spe | New 4-parameter Cut-off (Interpolated) | Sen/Spe |
| --- | --- | --- | --- | --- | --- | --- |
| Age | 59.61 | 0.59/0.64 | 59.67 | 0.59/0.64 | 59.82 | 0.59/0.64 |
| BMI | 25.05 | 0.54/0.53 | 25.04 | 0.54/0.53 | 24.99 | 0.54/0.52 |
| Nodule Diameter | 14.65 | 0.75/0.78 | 14.77 | 0.75/0.78 | 14.45 | 0.75/0.78 |
| CT Value | -436.14 | 0.79/0.83 | -438.99 | 0.79/0.82 | -442.81 | 0.8/0.81 |
| CTR | 0.18 | 0.85/0.8 | 0.22 | 0.83/0.84 | 0.20 | 0.84/0.83 |
| CEA | 2.37 | 0.56/0.64 | 2.39 | 0.54/0.65 | 2.31 | 0.59/0.62 |
| CYFRA21-1 | 2.01 | 0.6/0.54 | 2.02 | 0.59/0.55 | 1.98 | 0.61/0.53 |
| SCC | 0.99 | 0.4/0.46 | 1 | 0.39/0.47 | 0.97 | 0.4/0.46 |
| ProGRP | 42.8 | 0.4/0.61 | 42.83 | 0.4/0.61 | 42.52 | 0.41/0.6 |
| NSE | 19.63 | 0.34/0.62 | 19.74 | 0.34/0.63 | 19.47 | 0.35/0.60 |

**Table 4:** Cut-off points, sensitivity/specificity in the training and testing sets for HCC. When only one Sen/Spe value is shown, it is because sensitivity and specificity have the same value.

| Biomarker | Model | Cut-off Point | Sen/Spe (Training) | Sen/Spe (Testing) |
| --- | --- | --- | --- | --- |
| AFP | Empirical | 19.97 | 0.628 | 0.61/0.81 |
|  | Logistic (2P) | 8.41 | 0.77 | 0.69/0.6 |
|  | Logistic (4P) | 8.46 | 0.78 | 0.69/0.6 |
|  | ThresholdXpert 1.0 | 8.97 | 0.74/0.79 | 0.67/0.6 |
| CTP | Empirical | 5.75 | 0.51 | 0.47/0.67 |
|  | Logistic (2P) | 5.83 | 0.51 | 0.47/0.67 |
|  | Logistic (4P) | 5.99 | 0.40 | 0.47/0.67 |
|  | ThresholdXpert 1.0 | 5.47 | 0.35/0.68 | 0.47/0.67 |
| DKK-1 | Empirical | 391.9 | 0.6 | 0.61/0.70 |
|  | Logistic (2P) | 411.33 | 0.62 | 0.59/0.77 |
|  | Logistic (4P) | 399.6 | 0.61 | 0.61/0.74 |
|  | ThresholdXpert 1.0 | 389.56 | 0.6/0.6 | 0.61/0.70 |
| AGE | Empirical | 58.92 | 0.56 | 0.59/0.58 |
|  | Logistic (2P) | 59.54 | 0.55 | 0.53/0.58 |
|  | Logistic (4P) | 59.44 | 0.55 | 0.53/0.58 |
|  | ThresholdXpert 1.0 | 58.46 | 0.56/0.56 | 0.59/0.58 |
| MELD | Empirical | 6.92 | 0.35 | 0.41/0.23 |
|  | Logistic (2P) | 6.8 | 0.35 | 0.42/0.23 |
|  | Logistic (4P) | 6.77 | 0.35 | 0.42/0.23 |
|  | ThresholdXpert 1.0 | 6.87 | 0.35/0.35 | 0.41/0.23 |
| OPN | Empirical | 80.22 | 0.63 | 0.58/0.61 |
|  | Logistic (2P) | 82.92 | 0.66 | 0.55/0.61 |
|  | Logistic (4P) | 81.15 | 0.64 | 0.58/0.61 |
|  | ThresholdXpert 1.0 | 81.29 | 0.63/0.65 | 0.56/0.61 |
| PIVKA-II | Empirical | 4.12 | 0.68 | 0.59/0.67 |
|  | Logistic (2P) | 5.11 | 0.72 | 0.56/0.75 |
|  | Logistic (4P) | 4.79 | 0.69 | 0.56/0.75 |
|  | ThresholdXpert 1.0 | 4.45 | 0.67/0.65 | 0.58/0.68 |

### 3.2. Application in Hepatocellular Carcinoma: Individual Characterization and Comparative Model Analysis

Once the robustness of the methodology was verified in the pulmonary dataset, a detailed analysis of the hepatocellular carcinoma (HCC) training set was performed. In this critical phase, the following seven variables were individually characterized (AFP, PIVKA-II, OPN, DKK-1, Age, CTP, and MELD) using a comparative approach based on two distinct mathematical models: the standard two-parameter logistic model (2P) and the extended four-parameter logistic model (4P). In addition to these, the empirical calculation was also applied to determine the intersection point between the interpolated sensitivity and specificity data series.

The implementation of this approach responds to the need to capture the biological complexity of tumor biomarkers. While the 2P model provides a robust approximation assuming an ideal sigmoidal distribution (normalized transition from 0 to 1), the 4P model introduces additional scaling parameters (lower asymptote and maximum amplitude), allowing more accurate fitting of asymmetric curves or those exhibiting elevated basal levels of biological “noise.” Therefore, the objective is not to replace the classical model, but to provide an additional analytical tool that avoids excessive simplification of data in biomarkers with complex nonlinear behavior. Both models were optimized using the Dual Simulated Annealing algorithm to ensure that the derived cut-off points correspond to global mathematical optima rather than local fitting artifacts [7].

### 3.3. Variable Selection and Panel Evaluation

When running ThresholdXpert 1.0 with all available variables, it was observed that the inclusion of certain clinical variables did not provide a significant incremental benefit to panel performance. In particular, variables such as Age, CTP Score, PIVKA-II, and DKK-1 showed high variability in their optimal thresholds across different algorithm runs (marked with an asterisk in Table 5), suggesting that their contribution to the model is unstable compared with other variables. However, in other panels, the calculated values became stable.

**Table 5:** Evaluation of candidate diagnostic panels using combinatorial optimization in the training set and validation in the test set. Cut-off points represent the mean value after 10 stochastic optimization iterations.

| Combination | Cut-off Point | Sen/Esp (Training) | Sen/Esp (Test) |
| --- | --- | --- | --- |
| AGE + CTP SCORE + inverse MELD<br>+ AFP + PIVKA-II + OPN + DKK-1 | *AGE: 134,43<br>*CTP SCORE: 234,17<br>inverse MELD: 0,25<br>AFP: 19,69<br>*PIVKA-II: 324,98<br>OPN: 180,14<br>*DKK-1: 1588,52 | 0.79/0.80 | 0.75/0.70 |
| inverse MELD + AFP + PIVKA-II +<br>OPN + DKK-1 | inverse MELD: 0,53<br>AFP: 24,97<br>PIVKA-II: 19,42<br>*OPN: 266,31<br>DKK-1: 612,62 | 0.77/0.80 | 0.73/0.75 |
| AFP + PIVKA-II + OPN + DKK-1 | AFP 19,76<br>PIVKA-II 19,76<br>OPN 172,39<br>DKK-1 612,32 | 0.77/0.79 | 0.77/0.72 |
| AFP + PIVKA-II + inverse MELD | AFP: 11,12<br>*PIVKA-II: 59,45<br>inverse MELD: 0,25 | 0.79/0.77 | 0.75/0.6 |
| inverse MELD + AFP + OPN | inverse MELD: 0,25<br>AFP: 19,59<br>OPN: 171,55 | 0.79/0.80 | 0.75/0.68 |
| AFP + PIVKA-II + DKK-1 | AFP: 22,29<br>PIVKA-II: 19,79<br>DKK-1: 507,54 | 0.77/0.76 | 0.81/0.67 |
| AFP + inverse MELD | AFP: 11,09<br>inverse MELD: 0,26 | 0.78/0.78 | 0.73/0.6 |
| AFP + DKK-1 | AFP: 22,69<br>DKK-1: 505,04 | 0.76/0.76 | 0.81/0.67 |
| AFP + PIVKA-II | AFP: 8,97<br>PIVKA-II: 19,73 | 0.75/0.79 | 0.67/0.6 |

**Table 6:** Diagnostic Performance of Combined Panels (Validation in Test Set)

| Panel | Combination | Sensitivity (Test) | Specificity (Test) | Clinical Interpretation |
| --- | --- | --- | --- | --- |
| **Panel A** | AFP + PIVKA-II + OPN + DKK-1 | **0.77** | **0.72** | Replicates and validates the promising results reported in the literature. |
| **Panel B** | AFP + DKK-1 | **0.81** | 0.67 | Maximum Sensitivity. Ideal for surveillance in high-risk populations where no cases can be missed. |
| **Panel C** | AFP + PIVKA-II + OPN + DKK-1 + **inverse MELD** | 0.73 | **0.75** | Maximum Specificity. The inclusion of inverse MELD filters false positives, providing the best overall balance. |
| *Ref. Jang et al. [2]* | AFP + DKK-1 | 0.78 * | 0.73 * | * Results reported only in the training set.* |

The final selection of panels was oriented toward those combinations that demonstrated greater diagnostic robustness and parsimony, allowing the best balances between sensitivity and specificity to be achieved in the test set.

The asterisk corresponds to cut-off points in which strong fluctuations were observed when calculated with ThresholdXpert 1.0. The reported cut-off value corresponds, as in the other cases, to the mean of 10 obtained values.

### 3.4. Multimarker Panel Performance and External Validation

Following variable selection, three strategic panels were configured whose diagnostic performance was validated in an independent test set. Unlike previous reference studies in this same cohort—which determined diagnostic performance globally—this methodology implements a strict separation between training and validation data. This approach ensures greater robustness and generalizability of the results, mitigating the risk of overfitting and providing evidence of external validity that strengthens the clinical utility of the proposed panels.

**Panel B** achieved an unprecedented sensitivity of 81.3% in validation. **Panel C**, by incorporating inverse MELD, slightly sacrificed sensitivity in favor of higher specificity (75.4%), confirming that preserved liver function is a powerful discriminator of HCC [6].

## 4. Discussion

The update of Stage 1 has strengthened the ability of the methodology to operate with complex biomedical data. The initial validation in lung nodules demonstrated that the improvements produce results in a more automated manner, being similar and in some cases superior. In the case of individual analysis for CTR, both the 2-parameter and 4-parameter logistic fits yielded results similar to those obtained with the empirical cut-off and those obtained using ThresholdXpert 1.0, providing greater consistency to the analysis.

In the context of HCC, the results were obtained by dividing the dataset into two sets (training and test), which provides stronger evidence while avoiding overfitting. This was also done in the case of lung cancer.

A central finding is the role of inverse MELD (1/MELD) in Panel C. Low MELD values were consistently identified as a predictor of HCC in this cohort. This was interpreted as an “inflammatory noise” filter: patients with decompensated cirrhosis (high MELD) often show nonspecific AFP elevation. By filtering these cases using inverse MELD, the algorithm calculates the combination of cut-offs that best detects the true tumor signal, achieving the most balanced panel of the study.

It is worth mentioning that in the empirical calculation of the individual AFP cut-off, a considerably higher value (19.97) was obtained compared with the others (8.41–8.97), resulting in lower performance. However, in general, the four calculation methods tend to produce similar values. This reinforces the idea that cut-off points should be calculated using different approaches to rule out anomalies.

## 5. Conclusion

This study demonstrates that the performance of a multimarker diagnostic panel depends not only on the biological selection of biomarkers, but also on the mathematical robustness of the framework used to characterize and combine them.

The integration of empirical calculation, globally optimized logistic modeling using Dual Simulated Annealing, and independent combinatorial optimization within ThresholdXpert 1.0 constitutes a coherent and robust framework for the objective determination of cut-off points and the design of parsimonious panels.

Methodological validation in the pulmonary nodule dataset confirmed the stability and reproducibility of the approach, while its application to hepatocellular carcinoma allowed the identification of an optimized panel incorporating the inverse MELD variable, improving diagnostic balance in independent external validation compared with exclusively molecular panels.

This finding demonstrates that proper mathematical integration not only enables optimization of existing biomarkers, but also reveals relevant clinical-biological interactions, facilitating discrimination between tumor signal and underlying pathophysiological alterations.

Taken together, these results establish this framework as a generalizable tool for the rational development of binary diagnostic systems, with potential application across multiple biomedical contexts.

## 6. Data Availability

The datasets analyzed in this study are publicly accessible and available in their original publications. Complete references and corresponding links are included in the References section. No new experimental data were generated in this work.

## 7. Code Availability

ThresholdXpert 1.0 is available as free, open-source software (GNU GPL v.3 license). The complete source code, along with the documentation, can be accessed at:

https://github.com/roberto117343/ThresholdXpert

The program and scripts allow for the full reproduction of the results presented in this study.

## 9. Appendices

### Mathematical Algorithm Update in Python

In this study, new Python scripts have been implemented to act as the calculation engine for Stage 1.

#### 9.1. Script for Calculating the Empirical Intersection (Interpolation)

**Description:** It uses interpolation to mathematically determine the exact cut-off point where the sensitivity and specificity curves intersect, eliminating the subjectivity of visual inspection.

**GitHub Address:** https://github.com/roberto117343/ThresholdXpert/blob/main/Scripts/S3.1%20new.py

*#* «Copyright 2026 Roberto Reinosa Fernández» #\**

*#* This program is free software: you can redistribute it and/or modify*

*# it under the terms of the GNU General Public License as published by*

*# the Free Software Foundation, either version 3 of the License, or*

*# (at your option) any later version*.

*# This program is distributed in the hope that it will be useful*,

*# but WITHOUT ANY WARRANTY; without even the implied warranty of*

*# MERCHANTABILITY or FITNESS FOR A PARTICULAR PURPOSE. See the*

*# GNU General Public License for more details*.

*# You should have received a copy of the GNU General Public License*

*# along with this program. If not, see <http://www.gnu.org/licenses/>*.

*#\**

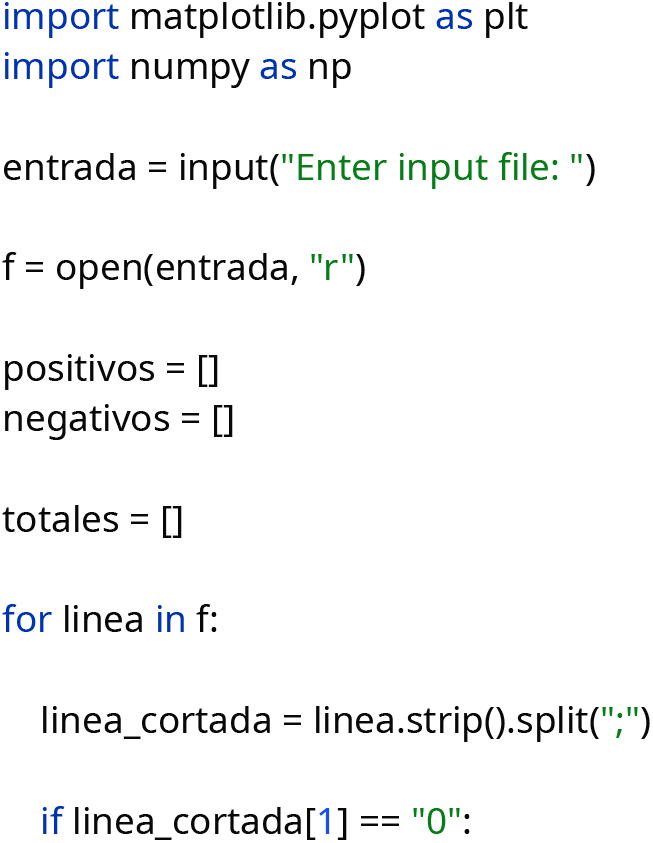
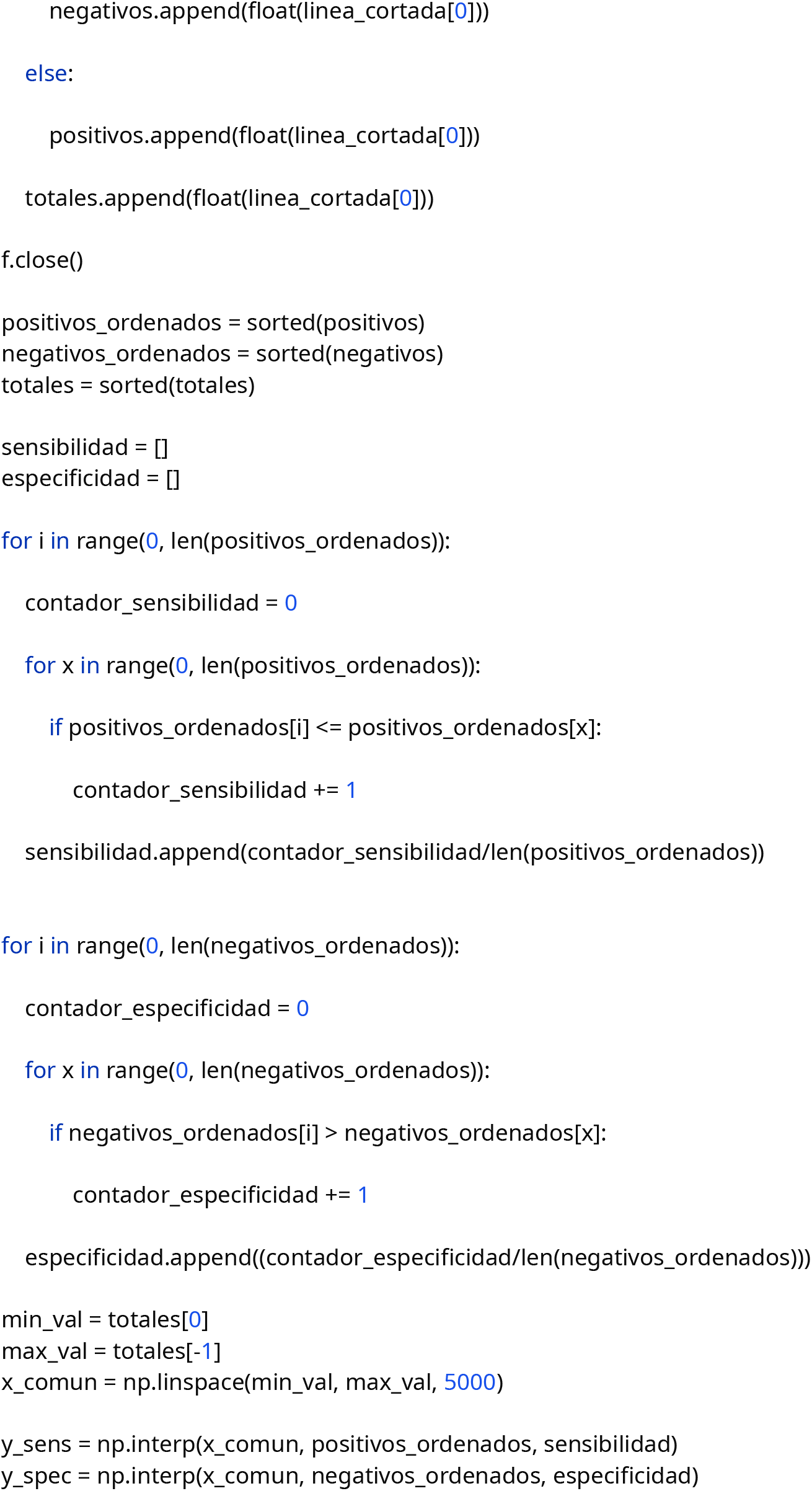
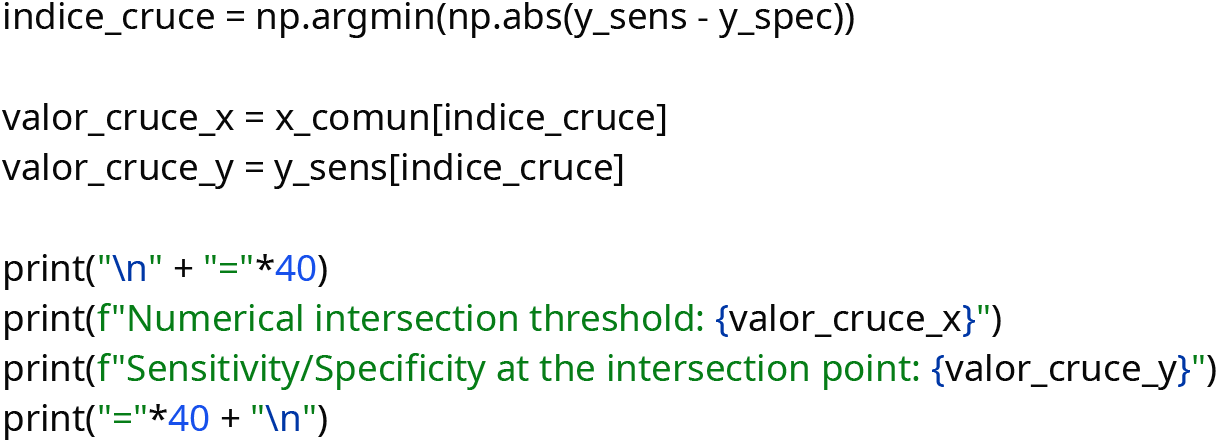

#### 9.2. Script for Fitting Two-Parameter Logistic Models using Dual Annealing

**Description:** It implements the dual annealing global optimization algorithm to fit 2-parameter models.

**GitHub Address:** https://github.com/roberto117343/ThresholdXpert/blob/main/Scripts/2P.py

*#* «Copyright 2026 Roberto Reinosa Fernández»*

*#\**

*#* This program is free software: you can redistribute it and/or modify*

*# it under the terms of the GNU General Public License as published by*

*# the Free Software Foundation, either version 3 of the License, or*

*# (at your option) any later version*.

*# This program is distributed in the hope that it will be useful*,

*# but WITHOUT ANY WARRANTY; without even the implied warranty of*

*# MERCHANTABILITY or FITNESS FOR A PARTICULAR PURPOSE. See the*

*# GNU General Public License for more details*.

*# You should have received a copy of the GNU General Public License*

*# along with this program. If not, see <http://www.gnu.org/licenses/>*.

*#\**

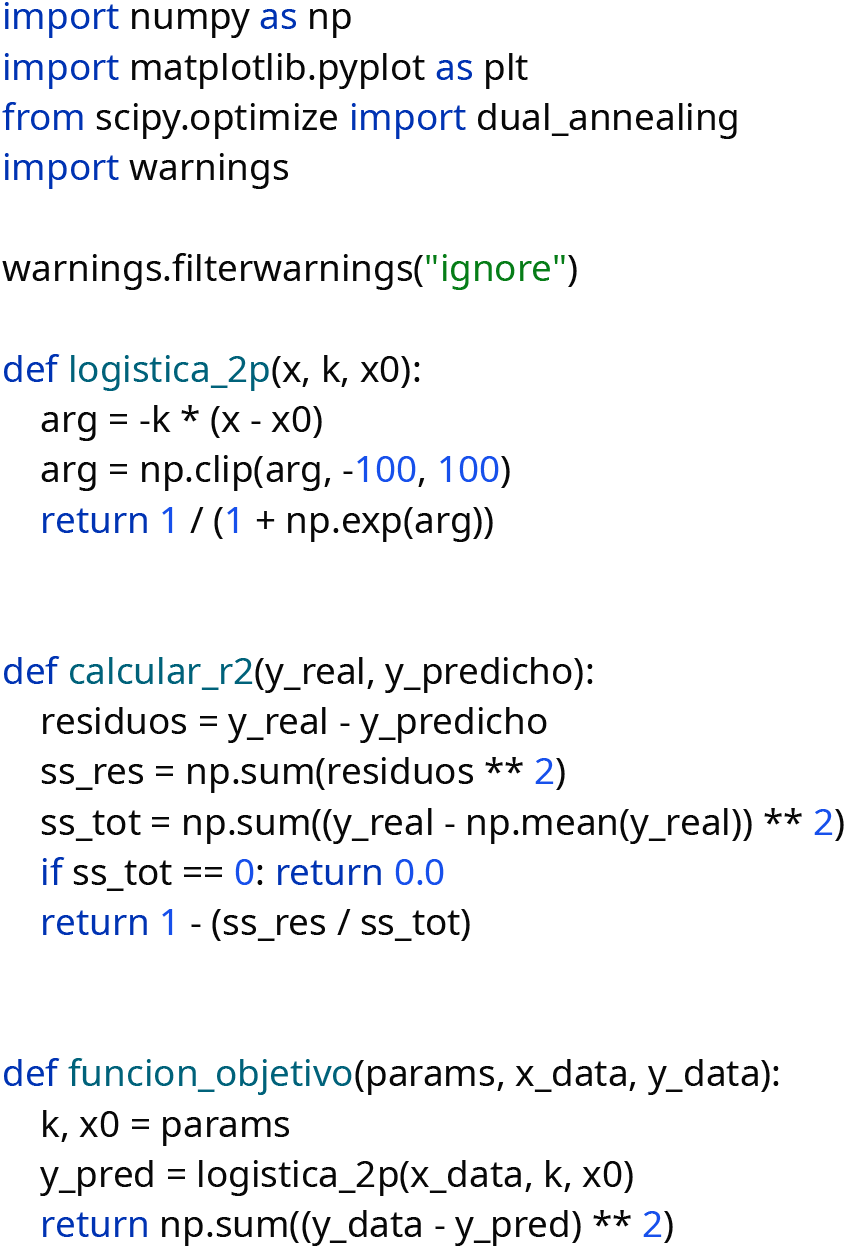
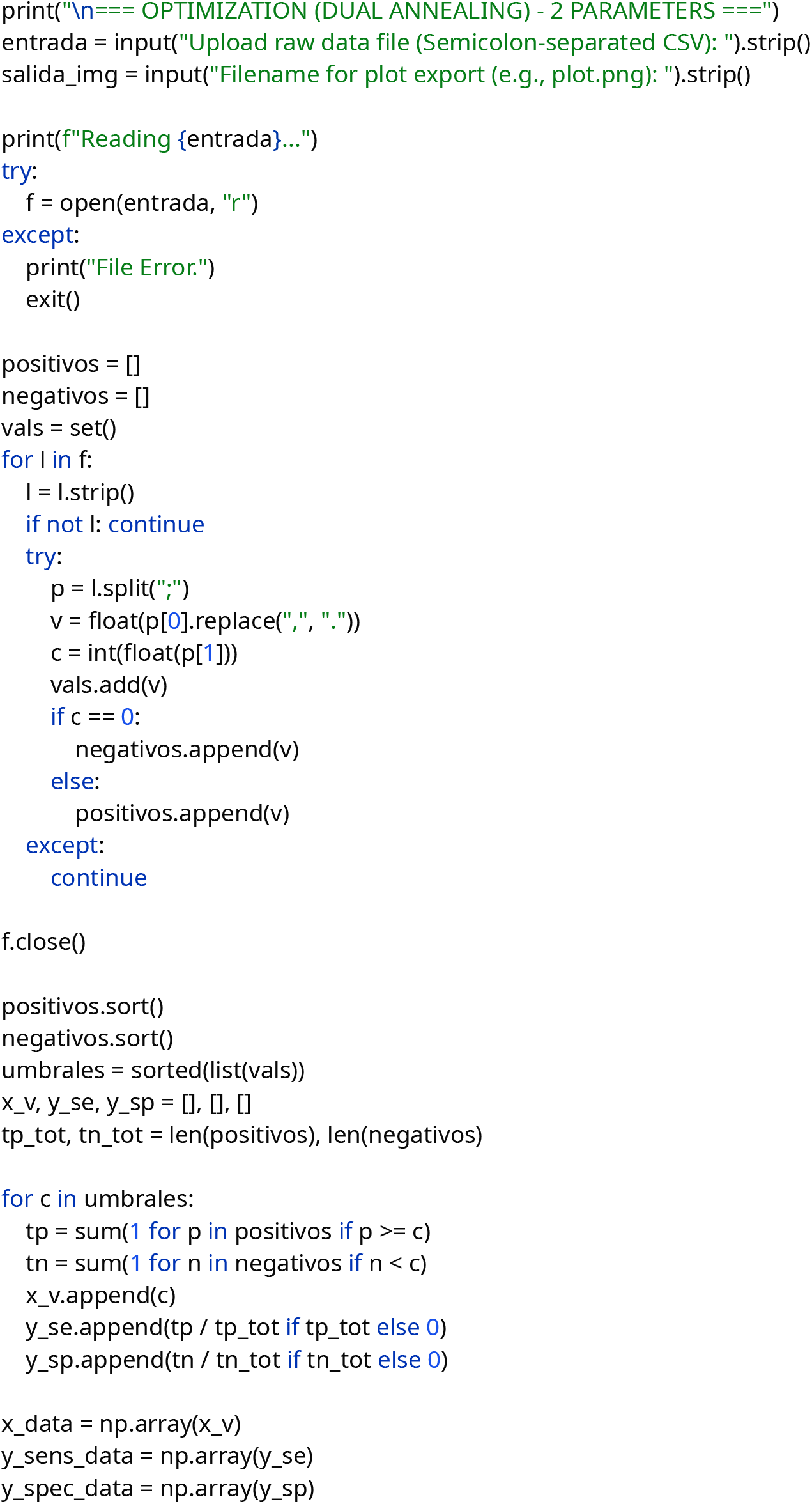
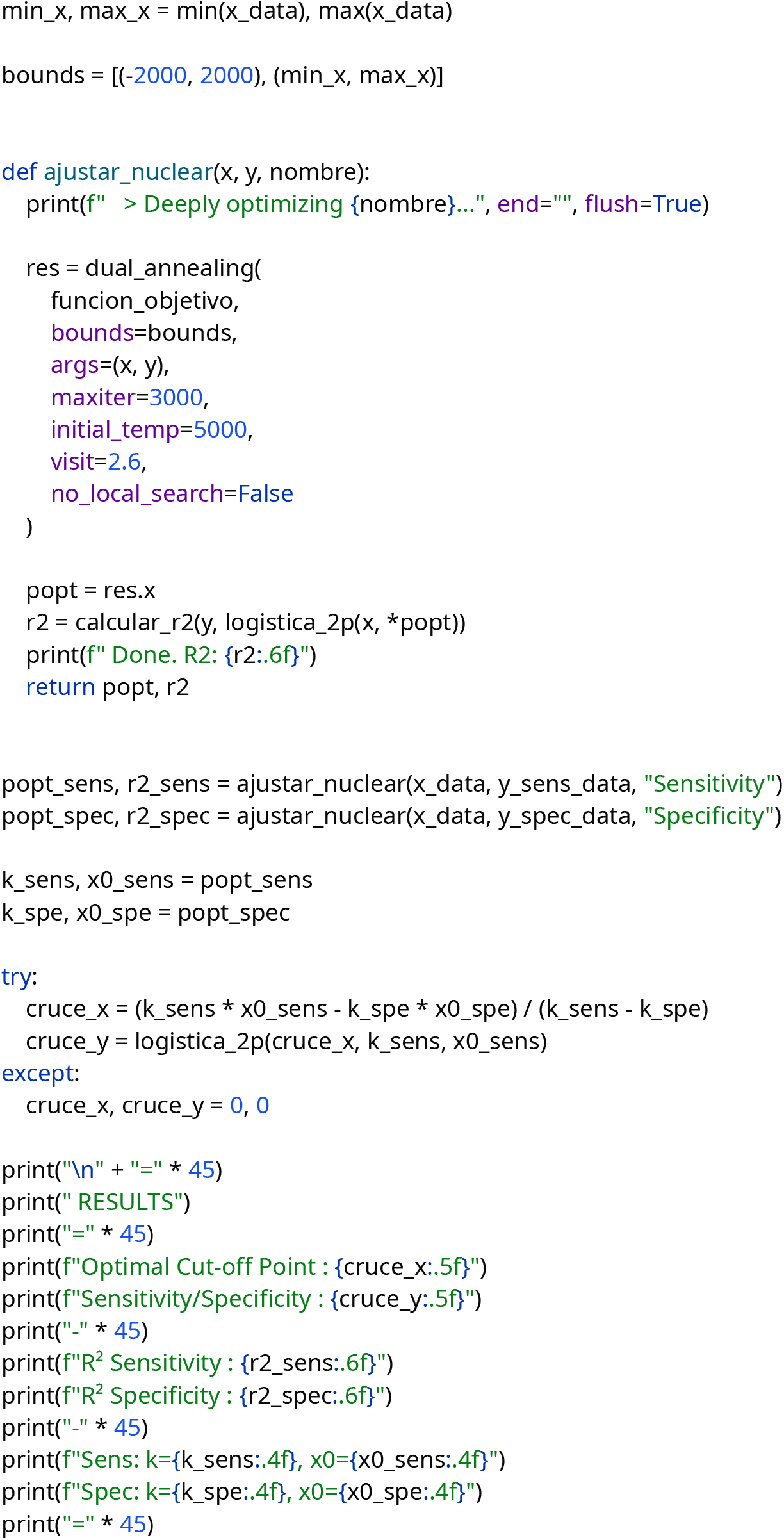
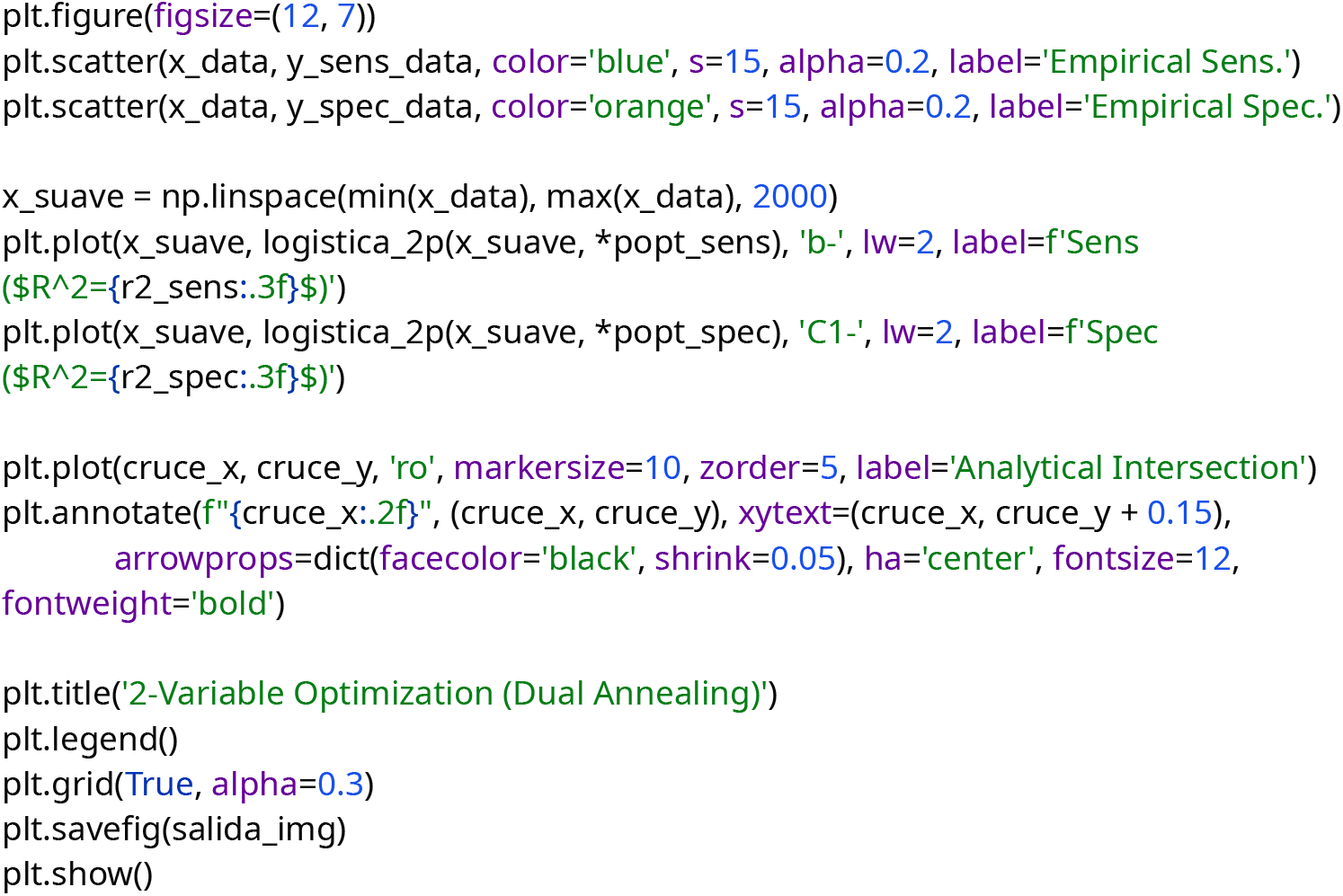

#### 9.3. Script for Fitting Four-Parameter Logistic Models using Dual Annealing

**Description:** It implements the dual annealing global optimization algorithm to fit 4-parameter models.

**GitHub Address:** https://github.com/roberto117343/ThresholdXpert/blob/main/Scripts/4P.py

*#* «Copyright 2026 Roberto Reinosa Fernández» #\**

*#* This program is free software: you can redistribute it and/or modify # it under the terms of the GNU General Public License as published by # the Free Software Foundation, either version 3 of the License, or*

*# (at your option) any later version*.

*# This program is distributed in the hope that it will be useful*,

*# but WITHOUT ANY WARRANTY; without even the implied warranty of # MERCHANTABILITY or FITNESS FOR A PARTICULAR PURPOSE. See the*

*# GNU General Public License for more details*.

*# You should have received a copy of the GNU General Public License # along with this program. If not, see <http://www.gnu.org/licenses/>. #\**

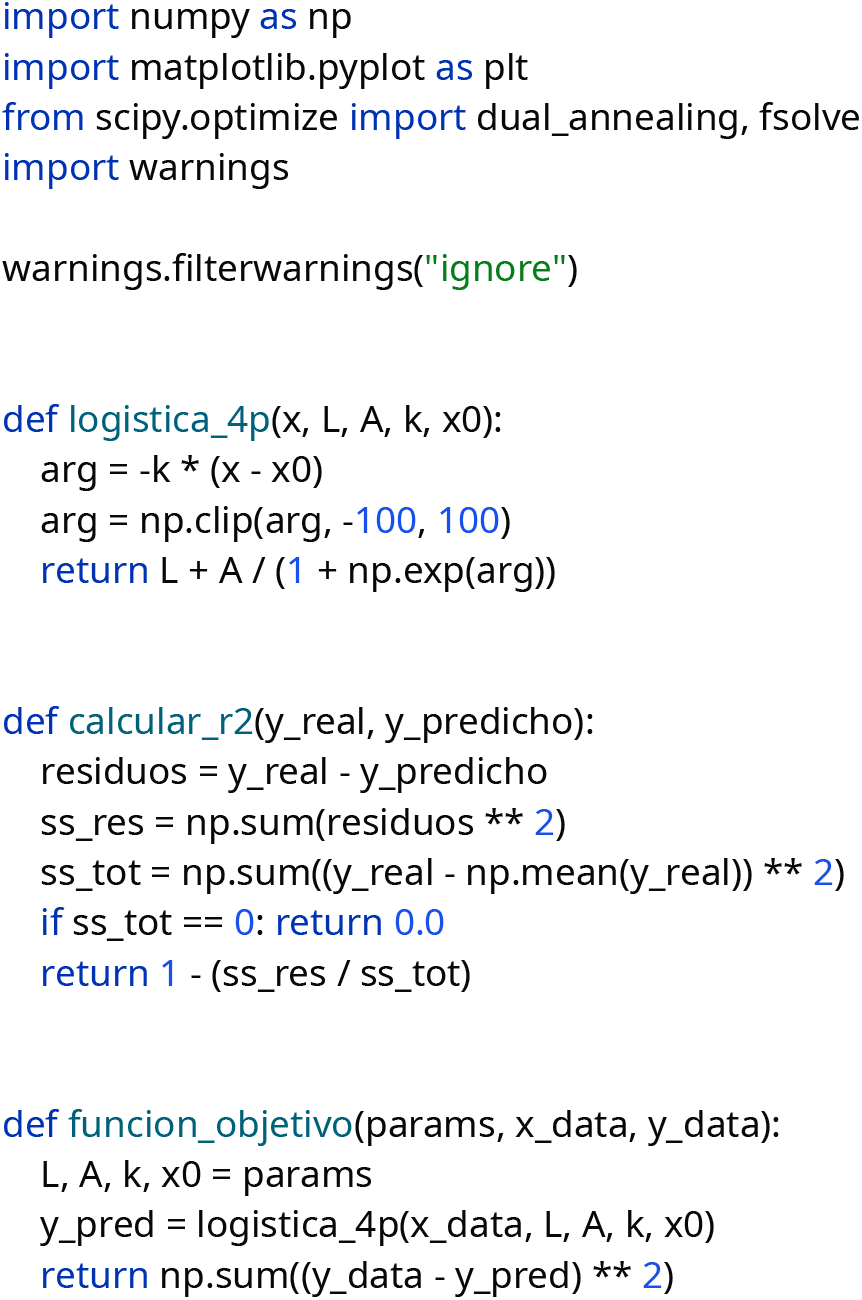
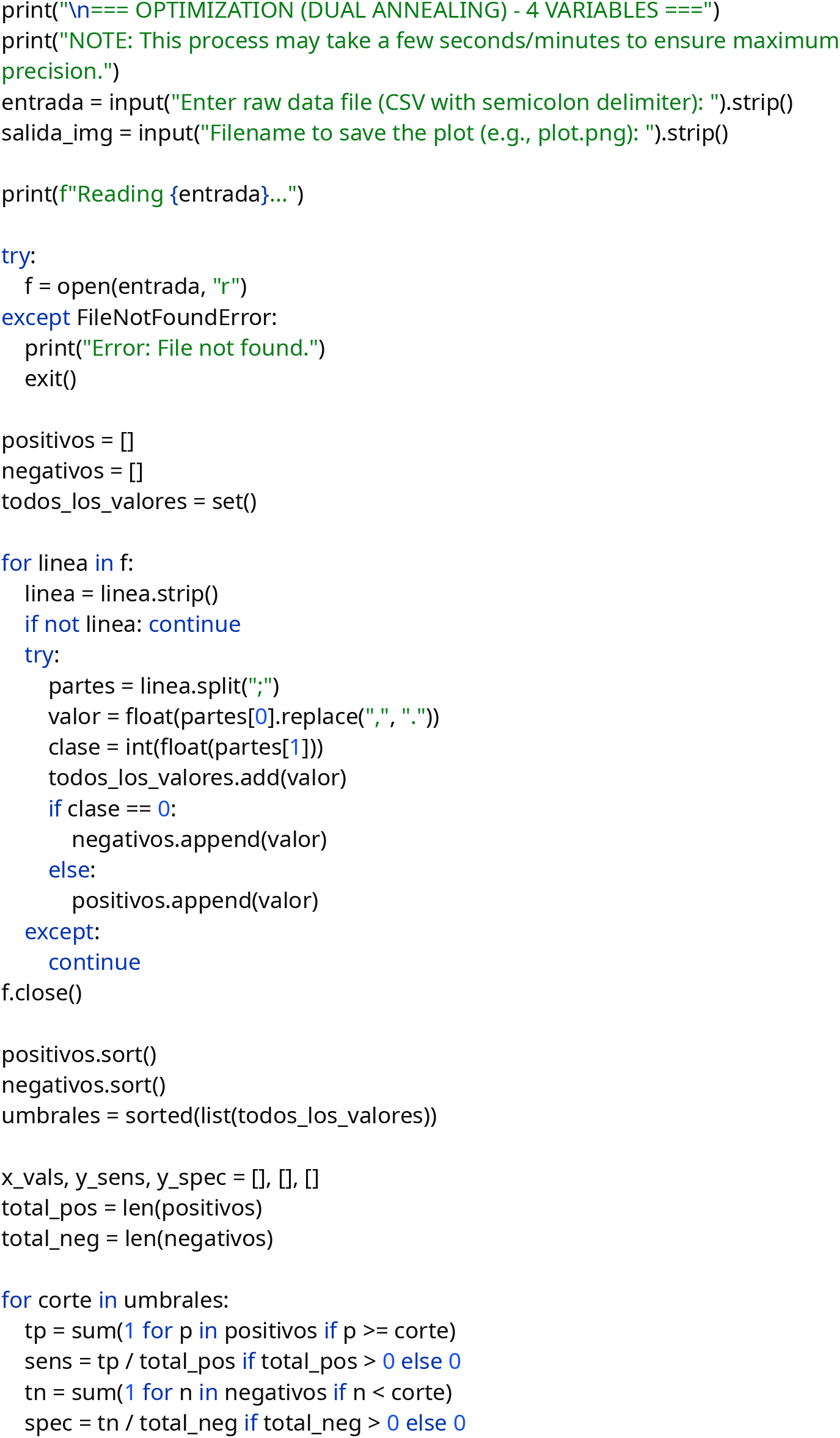
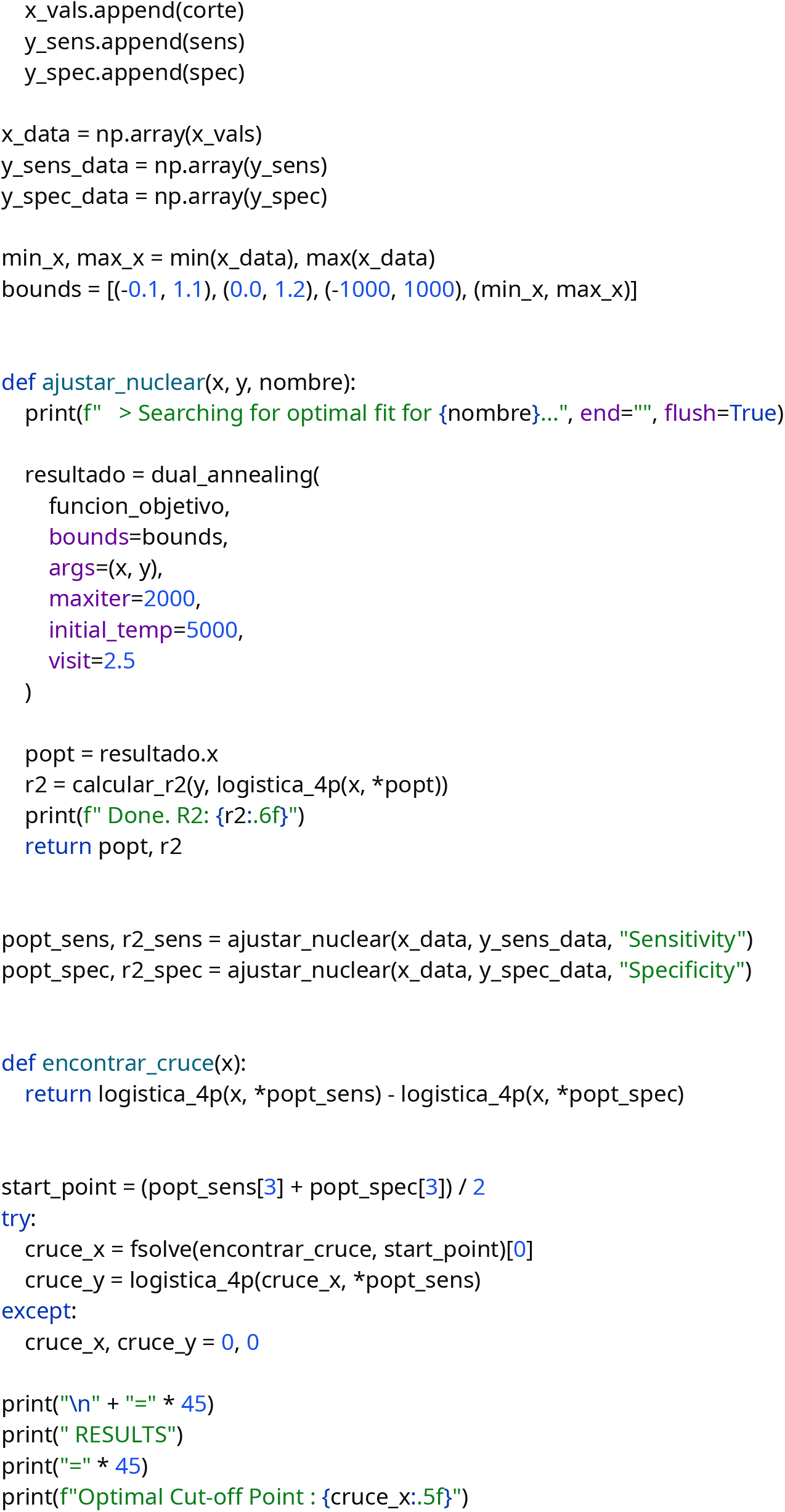
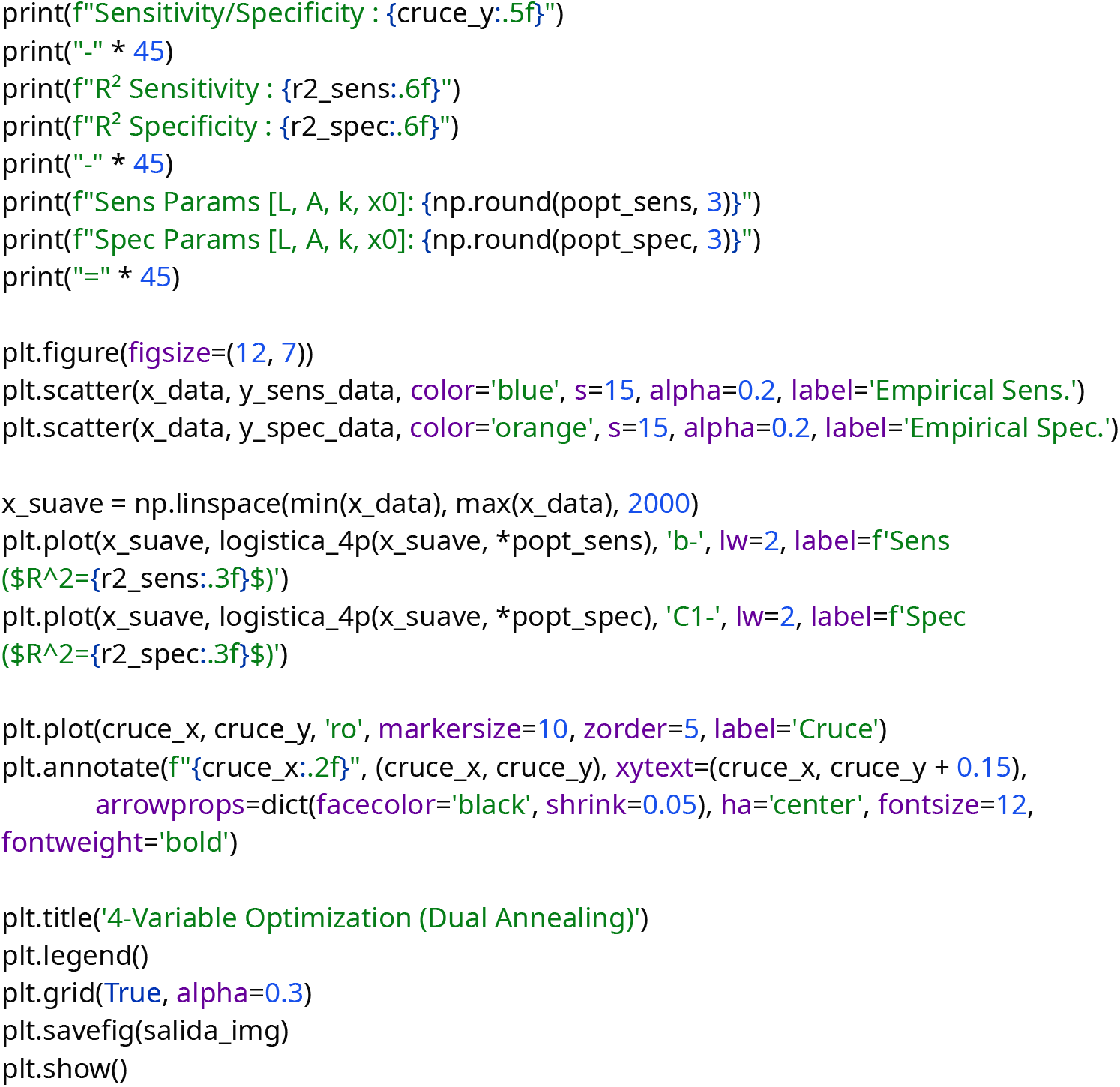

#### 9.4 Table of Variables, Definitions, and Units from the Original Datasets

This table summarizes the clinical, radiological, and biomarker variables used (Pulmonary Nodules and Hepatocellular Carcinoma).

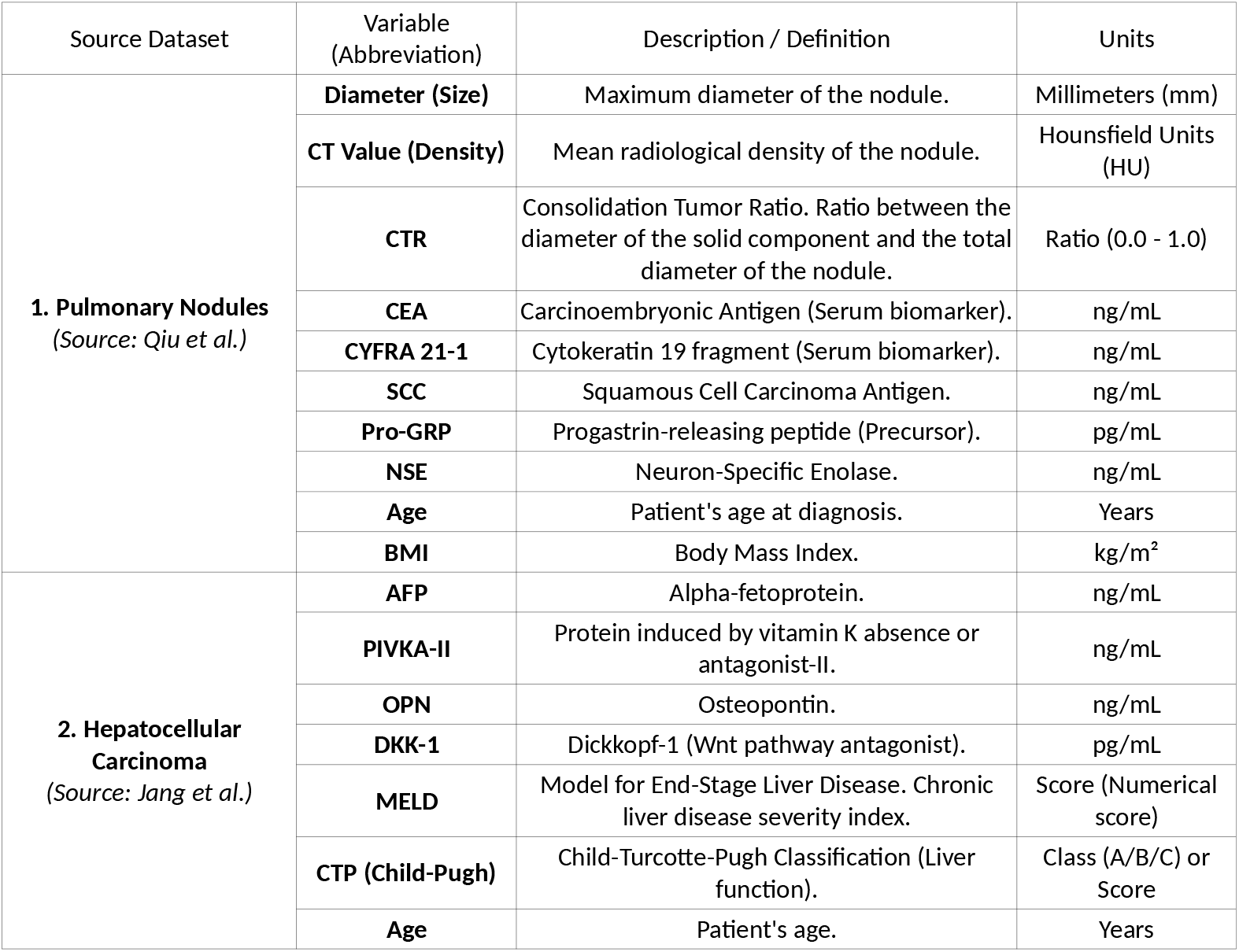

